# LLMs for analyzing open text in global health surveys: why children are not accessing vaccine services in DRC

**DOI:** 10.1101/2024.11.14.24317253

**Authors:** Roy Burstein, Eric Mafuta, Joshua L. Proctor

## Abstract

This study evaluates the use of large language models (LLMs) to analyze free-text responses from large-scale global health surveys, using data from the Enquête de Couverture Vaccinale (ECV) household coverage surveys from 2020, 2021, 2022, and 2023 as a case study. We tested several LLM approaches varying from zero-shot and few-shot prompting, fine-tuning, and a natural language processing approach using semantic embeddings to analyze responses on reasons caregivers did not vaccinate their children. Performance ranged from 61.5% to 96% based on testing against a curated benchmarking dataset drawn from the ECV surveys, with accuracy improving when LLM models were fine-tuned or provided examples for few-shot learning. We show that even with as few as 20–100 examples, LLMs can achieve high accuracy in categorizing free-text responses. This approach offers significant opportunities for reanalyzing existing datasets and designing surveys with more open-ended questions, providing a scalable, cost-effective solution for global health organizations. Despite challenges with closed-source models and computational costs, the study underscores LLMs’ potential to enhance data analysis and inform global health policy.

## Introduction

Generative artificial intelligence (AI) tools, particularly large language models (LLMs) like OpenAI’s GPT-4, are poised to transform a variety of fields, including Global Health. The ability to make informed, evidence-based decisions is critical in Global Health to optimize interventions, responsibly allocate funding, and maximize positive outcomes for populations. Vaccination programs targeting preventable childhood diseases exemplify this need, especially since vaccines are highly effective, safe, and cost-efficient. According to the World Health Organization, vaccinations save 2-3 million every year by protecting children from diseases like diphtheria, tetanus, whooping cough, and measles [1]. However, an estimated 14.5 million children were missing any vaccination in 2023; these children are colloquially referred to as zero-dose children [15]. Understanding the various barriers to vaccination for these zero-dose children, particularly through large-scale, nationally representative data collection efforts, remains a challenge for the Global Health community [2]. In this article, we explore how LLMs can enhance the analysis of qualitative data from global health national surveys, offering a powerful new tool to extract insights from free-text responses at scale about reasons why children are not accessing vaccine services.

One traditional avenue for generating evidence for Global Health decision-making is through the implementation and use of large-scale, nationally representative surveys that gather structured data—numerical, categorical, or ordinal—about individuals, families, and communities [3]. These data can be used to estimate key population health indicators at the national or subnational geographic resolution [1,4]. Alternatively, unstructured qualitative data collected in smaller scale surveys or through targeted interviews offer more detailed and nuanced information [5]; however, such data can be challenging to analyze at scale, requiring considerable time and expertise to code and analyze [6]. In this article, we demonstrate that LLMs, with a small amount of human researcher time, can analyze qualitative data in the form of free text responses from large-scale surveys with high fidelity, unlocking new insights from survey data. We demonstrate this approach by analyzing free-text response data from large-scale surveys called the Enquête de Couverture Vaccinale (ECV) household coverage surveys conducted in the Democratic Republic of Congo (DRC) in 2020, 2021, 2022, and 2023 which includes the option for free-text responses on the reasons caregivers provide for not vaccinating children. [12]

Analyzing free-text responses for topic modeling, summarization, and categorization has been a major focus of the natural language processing (NLP) community over the last decade. With new NLP techniques, transformer-based neural networks, and the recently released pre-trained, highly performant large language models (LLMs), the community has demonstrated substantial success on a variety of textual analysis tasks [7,8,9,10]. For example, OpenAI’s generative pre-trained model GPT-4 and other recently released LLMs can surpass human benchmarks on visual commonsense reasoning, multitask language understanding, and competition-level mathematics [11,13]. The broader scientific community is beginning to adopt these models for a large variety of tasks, including in the social sciences community with novel uses of topic modeling on survey data [25]. One of the more compelling elements of using these new LLMs is the zero-shot (does not requiring specialized training or data on a new task) performance of LLMs can be impressive, requiring only the definition of the LLM role and thus minimal hands-on time from researchers or artificial intelligence (AI) experts or teams. It has also become clear, though, that for a variety of tasks, providing the model with some additional input or context—known as prompt engineering, few-shot, or fine-tuning approaches—can significantly enhance performance, often without extensive cost in expertise and computational effort [12]. In this study, we implement a broad range of these methodologies and models to evaluate their performance, accuracy, implementation effort, and cost-effectiveness in analyzing the data from surveys.

In this article, we analyze the free response text from the ECV surveys on reasons for non-vaccination using large language models (LLMs). We begin by describing the underlying dataset and the process for creating a benchmark validation set within the response dataset. Following this, we outline how different approaches and underlying models can categorize these responses into both original and newly identified categories. We then show how these data provide substantive and programmatic insights not anticipated by the survey designers or implementors. We conclude with a discussion on how this approach can be easily generalized to other similar surveys and the broader applicability of this type of data analysis in the Global Health space.

## Methods

### Data Source

We used data from the 2020, 2021, and 2022 cross-sectional rounds of the Enquête de Couverture Vaccinale (ECV) household coverage surveys to test and the LLM-based categorization models. Survey respondents are caregivers of children 6-23 months old who report on their use and perceptions of vaccine services. The 2020 round was conducted in 18 provinces, and the 2021 and 2022 rounds were expanded to all 26 provinces in the country. Detailed descriptions of these surveys have been provided elsewhere [16,14,17] and a survey protocol is available in the project’s github repository (https://github.com/InstituteforDiseaseModeling/AIAugmentedSurveyResponseCategorization/tree/main).

In the 2021 and 2022 surveys, if children 12-23 months did not receive all recommended vaccines, their caregivers were asked the following questions in French: “vs89: *Pourquoi l’enfant n’a-t-il pas reçu tous les vaccins recommandés? vs102: Parmi les raisons précédentes, quelle est la raison la plus importante?”*, translated as: “vs89: *Why has the child not received all the recommended vaccines? vs102: Among the previous reasons, which is the most important one?”*. For the ‘most important’ question, there were 22 response options available, with a 23^rd^ option of ‘Other: free text”, see ***Figure 1*** for a full list of response options. In the 2020 survey only the following question was asked: *“qa401: Pourquoi l’enfant n’a pas été complètement vacciné(e)?”*, translated as: *“Why has the child not been fully vaccinated?”*. This question was multiple choice and allowed for an ‘Other (please specify)” option. Overall, the response options were similar, with some differences in wording, and with the exclusion of ‘Mother is too busy’ as an option, and the question was asked of caregivers with children 6-23 months old. Furthermore, the 2020 survey was only conducted in 18 of the 26 provinces of DRC, so it is not considered nationally representative, while all others are.

**Figure 1:**
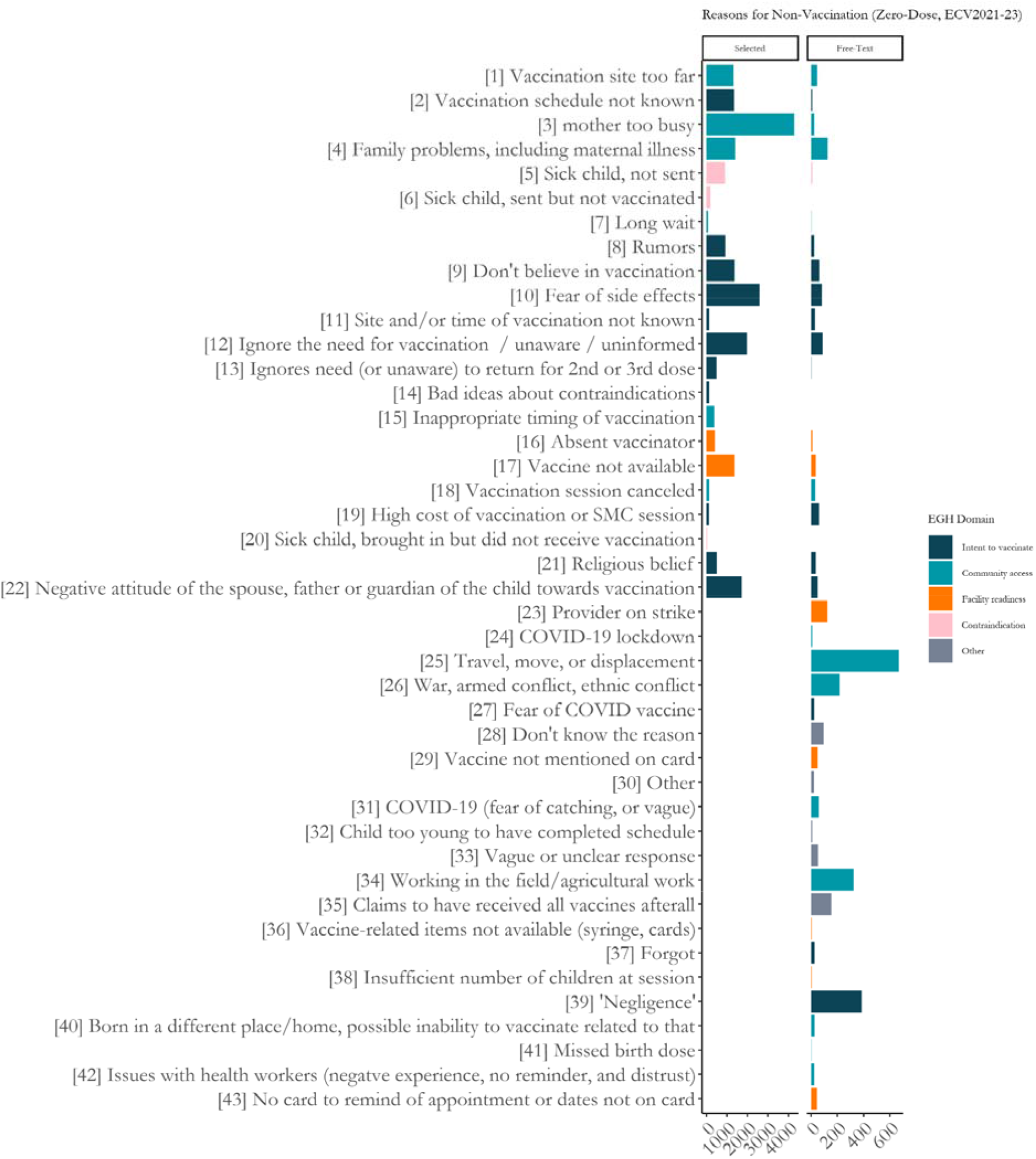
Reasons cited for non-vaccination among caregivers of zero-dose children in the 2021, 2022, and 2023 ECV Surveys. 2020 was excluded from this plot as that survey used a slightly different set of response options. The left panel shows the selected options, and the right panel shows the free-text responses categorized using the fine-tuned model.

### Creation of additional categories

Given that the respondent selected ‘Other’ it was our assumption that the original set of 23 categories would not sufficiently cover the set of reasons given in the free text, and as such we added new categories. We chose to pre-specify the full set of category options to ensure our set was collectively exhaustive. First, we included categories from the first question vs89, which were not available in vs102, the primary question of interest. Next, in creating the benchmarking dataset detailed in the following section, we evaluated a random sample of 1000 replies and created new distinct categories for any group of five or more similar responses. While we aimed for categories to be mutually exclusive, some categories where similar but with specific details that we posited would yield distinct insights. For example: “Travel, move, displacement” is a general category that has overlap with both “War, armed conflict, ethnic conflict”, as well as “Working in the field/agricultural work”. Finally, we included new categories for *Don’t know the reason, and Vague response*. A full list of the expanded set of categories is available in ***Figure 1***.

Alternative methods for category development could be employed by future researchers, such as leveraging LLMs to generate new categories either a priori or iteratively during the analysis. However, our initial experiments indicated that these approaches often produced an excessive number of overly specific categories or, in the case of pre-specification, omitted significant categories. To maintain control over this process and ensure the results remained aligned with policy-relevant insights, we adopted a human-in-the-loop, researcher-driven approach

### Benchmark dataset

Of the full set of 8807 unique responses from the 2020, 2021, and 2022 surveys, we randomly selected 999 to comprise a benchmark dataset. We first translated each response from French to English using GPT-4. Then, one of the authors (RB), read each response and assigned it to the best-fitting category. Of these, 799 responses were set aside for training and 200 for testing.

We assessed the AI-based categorization against the human researcher assigned category. As such, our approach does not represent a gold standard validation generalizable to human intelligence, but rather a practical benchmark to understand similarity of LLM-categorization relative to the human expert (author RB). To this end, we assessed accuracy for each method (number of responses the model categorized the same as the human divided by 200).

The test set size (200 samples) was chosen based on a power calculation with an assumed accuracy of 85%, 5% margin of error at a 95% confidence level 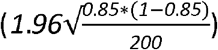. Given the large number of categories, 43 in all, and relatively small number of responses, we were not powered to assess category-specific accuracy.

### Choice of LLM

We selected GPT-4o (gpt-4-2024-08-06) as the foundation LLM for this study, based on its competitive performance across multiple benchmarks at the time of analysis. The focus of this work was not to compare different LLMs, but rather to demonstrate proof of concept that LLMs can effectively handle this survey analysis task. Given the rapid advancements in model development, we acknowledge that the specific accuracy metrics presented here are subject to change, as newer models continue to outperform previous iterations. Thus, our findings should be interpreted as indicative of the capabilities of LLMs in this context, rather than a definitive measure of performance.

### Categorization methods

We tested three classes of approaches to LLM-based free text categorization. First, a ‘Natural Language Processing (NLP)-like’ approach based on clustering semantic embeddings of response text, several direct prompting approaches for LLMs, and a fine-tuned LLM. The following subsections describe each in detail.

### “NLP”

We encoded each response using OpenAI’s text-embedding-3-small model, resulting in 1536-dimensional embedding vectors [18]. These embedding vectors are representations of the semantic meaning of each response, with the distance between them representing relatedness [19,20]. Therefore, we hypothesized that the survey responses that are closer in embedding space could represent the same category of reason for non-vaccination. We tested three clustering algorithms: k-means, gaussian mixture models (GMM), and hierarchical clustering with an increasing number of clusters, up to 250. GMM and K-means approaches yielded similar accuracies (61.5%) at 200 clusters; see ***Supplementary Figure 1*** for additional details. For comparison with the LLM-based approaches, we present the NLP results in this article using the GMM algorithm with 200 clusters.

Once each response was assigned to a cluster, we used the LLM to assign a category label to each cluster by comparing up to 25 randomly chosen example responses assigned to the given cluster with the full list of 43 categories. The following system role and prompt were used:

□ System Role: You are an assistant who looks at example responses to a survey question and describes their common thematic meaning. The survey was conducted in DRC and asked parents why their child did not receive all their recommended vaccines. I will provide you with a list of survey responses and a comprehensive list of thematic labels. You can only choose from one of these. You will only return the number associated with the chosen label, nothing else.
□ Prompt: Responses have been pre-screened and should all fit under one cohesive theme. Here are the example responses, separated by semicolons: [LIST OF UP TO 25 EXAMPLE RESPONSES HERE] Here is a list of possible labels separated by semicolons: [LIST OF ALL POSSIBLE REASON CATEGORIES HERE]

This approach has several outright advantages over the others, which are described below. First, this approach required lower resources as it only required the number of prompt completions commensurate to the number of clusters, rather than one prompt per response. At N clusters equal to the number of responses, this solution would converge on a zero-shot approach. Second, this approach is completely unsupervised, not requiring us to supply the model with response-category example pairs.

Our approach is similar to previous NLP techniques, such as BERTopic [26], but with a novel enhancement: we leverage an LLM to assign category labels to each cluster. To our knowledge, this is the first approach that combines unsupervised topic modeling with the LLM’s capacity for text summarization, while also integrating researcher intuition and domain expertise. We believe this method presents a promising future research direction for generating categories and topics in survey analysis, offering a more flexible and insightful approach to understanding free-text data and mitigating the sensitivity of solely using LLMs for topic discovery.

### Direct Prompting

We also tested several direct prompting approaches. These approaches represent the most straightforward way of utilizing a LLM for this task. Each of these involved showing the LLM a single response, all possible categories, and asking it to choose the category that best fits the response. We tested a zero-shot prompt, few-shot prompts with 20, 50, 400, and 799 training examples, and a chain of thought (CoT) prompts. Full prompts are available in the code files available in the project’s github repository: https://github.com/InstituteforDiseaseModeling/AIAugmentedSurveyResponseCategorization. Both zero-shot and CoT approaches are fully unsupervised.

### Fine-tuning

LLMs can be customized for specific use cases via fine-tuning [21]. We fine-tuned GPT-4o using the same 799 example response-category pairs we used in the few-shot learning approach. Once a model is fine-tuned, prompts can be much shorter, and thus lower per-prompt API costs after the initial investment in model training. For example, our fine-tuned model did not require the 799 examples to categorize each response which few-shot did, but it has higher upfront cost in terms of researcher time, compute resources, and complexity of implementation.

### Accuracy and Accuracy Ceiling

As described earlier in this section we used accuracy to assess the strict agreement between the human researcher and model-derived categorization. Though, there existed certain cases in which the model-derived categorization was arguably correct despite being discordant with the human-derived categorization. This occurred when the response was vague, when a response could fall into two not-quite-mutually-exclusive categories, or when there was a compound response that mentioned two categories. For example, a vague response like ‘Long journey’ could arguably yield a category choice ‘Vaccination site too far’, or ‘Travel, move, displacement’. To arrive at the adjusted ‘accuracy ceiling’, we reviewed all responses with discordant categorizations, and judged if the model-derived categorization was correct and included these in the numerator for the accuracy calculation.

### Survey analysis

After benchmark testing we applied the most successful method to the full survey data set. In addition to the 2020, 2021, and 2022 surveys, of which a subset was used to train and test, the 2023 survey was released after our benchmarking was conducted and was added to the collated dataset for this analysis step.

We ran the model for all unique responses and applied the results across the full set of responses. We calculated descriptive statistics, including the frequency of category assigned, the ratio of new categories to pre-existing ones, trends in category frequency over the three category years, and mapping the spatial distribution of some categories. These results and their insights were explored through data visualization. The 2020 survey is not included in ***Figure 1*** due to its difference in question format, lack of recency, and lack of national representativeness.

Raw survey responses to the free-text question and all code for this study in both python and R are available at https://github.com/InstituteforDiseaseModeling/AIAugmentedSurveyResponseCategorization

### Ethics Statement

The institutional review board of the Kinshasa School of Public Health Ethical committee gave ethical approval for the collection and use of survey data.

## Results

Of the 240,256 total responses across the 2020, 2021, and 2022 surveys, 94,663 caregivers were eligible for, and responded to this question, of those 17,940 chose the ‘Other’ option and gave a free-text response. Of those free text responses, 8807 were unique strings. The most common repeated matching responses were “Travel” (N=461), “Negligence” (N=291), “Vaccine schedule in progress” (N=232), and “Health worker strike” (N=232).

***Table 1*** lists all 8 LLM-based categorization approaches tested on the 200-response test set. Precise accuracy ranges from 61.5% for the NLP approach to 83.5% for the fine-tuned model with 799 examples. Generally, accuracy increases as researcher level-of-effort increases, with few-shot accuracy increasing as the number of training examples shown increases, though with marginal increases in accuracy declining after 50 examples, with a smaller difference with accuracy moving from 50 examples to 799 examples, than from zero to 50.

**Table 1:** Benchmarking and cost results for the LLM-categorization approaches tested.

| Approach | Precise Accuracy | Accuracy Ceiling | Upfront Cost (\$) | Avg. prompt tokens/ API call | Avg. completion tokens/ API call | Avg. Cost (\$) / API call | Estimated Total Cost for full dataset (\$) | Researcher level of effort |
| --- | --- | --- | --- | --- | --- | --- | --- | --- |
| NLP + Clustering (n=200) | 61.5% | 74.5% | 0.0003 | 605 | 1 | 0.0015 | 0.30 | Low (unsupervised) |
| GPT-4o Zero Shot | 71.5% | 87.0% | 0 | 398 | 1 | 0.0010 | 9.91 | Low (unsupervised) |
| GPT-4o Chain of Thought | 73.5% | 89.5% | 0 | 472 | 52 | 0.0017 | 16.77 | Low (unsupervised) |
| GPT-4o Few Shot (n = 20) | 76.0% | 93.0% | 0 | 621 | 1 | 0.0016 | 15.41 | Medium |
| GPT-4o Few Shot (n = 50) | 79.5% | 93.0% | 0 | 943 | 1 | 0.0024 | 23.36 | Medium |
| GPT-4o Few Shot (n=400) | 81.5% | 92.5% | 0 | 4486 | 1 | 0.0112 | 110.73 | High |
| GPT-4o Few Shot (n = 799) | 83.0% | 95.0% | 0 | 8464 | 1 | 0.0212 | 208.84 | Highest |
| GPT-4o Fine-tuned (n = 799) | 83.5% | 96.0% | 13 | 398 | 1 | 0.0010 | 22.91 | Highest |

Accuracy ceilings were between 11 and 16 percentage points higher than precise accuracy. 186 unique human- and model-categorized discordances were reviewed, with 74 assessed as having an acceptable model-derived categorization. At the highest, the fine tuned model achieved 96% and at the lowest the NLP approach achieved 74.5%. In certain cases, models were incorrectly categorized when responses appeared vague but contextual knowledge was needed to fully understand. For example, most models incorrectly categorizes “Vaccines give diseases to children” as *Fear of Side Effects*, while the researchers categorized this as *Rumors*, based on the understanding that there are rumors about disease causing agents being purposely placed in vaccines as a common rumor in DRC.

We estimated overall costs based on current OpenAI API costs as of September 2024: $2.5 per million input token and $10 per million output tokens [22]. Only the NLP approach and the fine-tuning approaches has upfront costs before asking the models to categorize responses. The NLP approach required producing embedding vectors for each response at a low overall cost of $0.0003 ($0.020 per million tokens), and the training cost for the fine-tuned model with 799 examples was $12.99 ($25 per million training tokens). Cost per API call was estimated using the average number of input and output tokens per method. Overall costs were by multiplying those costs for the 9865 unique string responses. Costs are lowest for the NLP approach because it only needs to be run once per cluster, in this case 200 times, versus once per unique response. For the few-shot responses, cost scale with number of examples given as these increase the number of input tokens per call, ranging from $9.91 in zero-shot up to $208.84 for the 799-shot. The fine-tuned model achieved similar accuracy to the 799-shot model with significantly lower overall cost ($22.91 versus $208.84) due to requiring significantly fewer input tokens.

Based on its superior cost-efficiency, we chose the fine-tuned model to apply back to the full set of survey responses. ***Figure 1*** shows the full set of categorized responses amongst zero-dose children in the 2021, 2022, and 2023 surveys. On the left panel are selected responses and on the right are the LLM-categorized ‘other’ responses. All categories starting at 23 were not included in the original vs102 survey question and are thus empty on the ‘selected’ side. Bars are colored according to their domain membership in the Exemplars in Global Health Vaccine Delivery Framework [23,24]. Among all the 24,870 zero-dose caregivers in these three surveys who responded to vs102, 12.3% responded *Other*, second only to *Mother too busy* (17.2%). Among those responding *Other*, 76% of the LLM-categorized responses were in the new categories. Newly categorized responses were more commonly around Community Access-related issues (51.0%) compared to Selected responses (34.7%). The most common three categories were *Travel, move, and displacement* (21.8%) and *‘Negligence’* (12.6%) and *Working in the field* (10.5%).

***Figure 2*** shows examples of categorized responses which vary in time. First, Covid-19-related reasons (*Fear of Covid-19 Vaccine; Covid-19 Lockdown;* and *Covid-19 (fear of catching or vague)*) peaked in 2021 with just 3.1% of other responses and has since reduced to almost zero. A major strike of healthcare workers happened in 2021 and was captured by responses peaking at 18.1% that year. Finally in the 2020 survey, which included children as young as 6-months, 37.8% of *other* respondents indicated that their child was too young to have completed their vaccination schedule. Indeed 96.2% of those responding ‘*Too Young’* were 9-months or younger – the age of the final scheduled vaccine, Measles, at the time of that survey. Interpretation of the 2020 requires caution as it was not nationally representative, though keeping only the 18 provinces in subsequent surveys yields similar trends.

**Figure 2:**
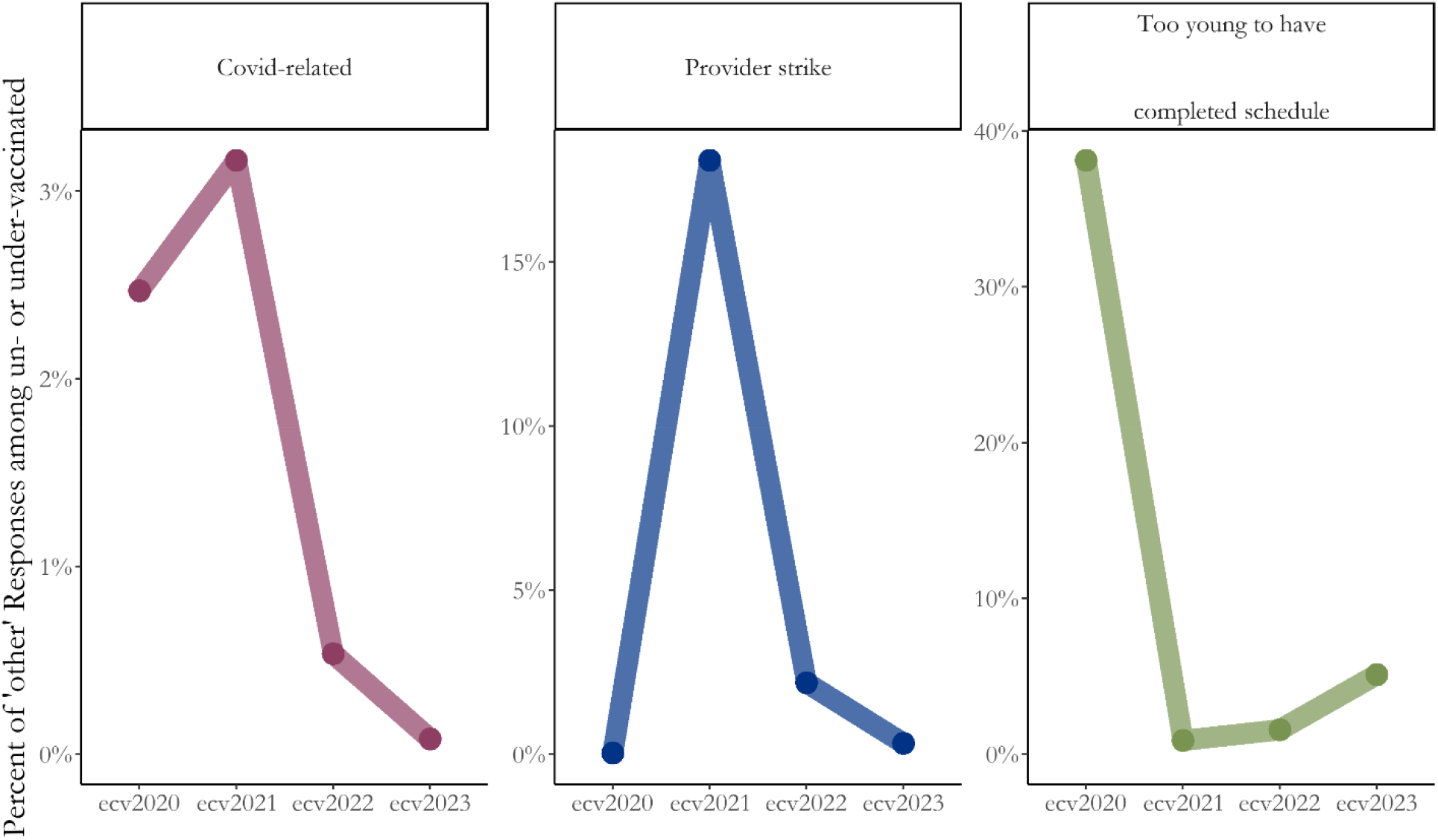
Proportion of categorized ‘other’ responses over time among caregivers of zero-dose and under-vaccinated children.

Linkage to other survey elements can yield additional insights. For example, in *Figure 3*, we show the percentage of caregivers with zero-dose or under-vaccinated children whose response was categorized as *War, armed conflict, ethnic conflict* as being heavily geographically concentrated in the east of the country, where conflicts are known to be ongoing. In some health areas, almost all respondents cite this as their primary barrier to accessing vaccine services for their children.

**Figure 3:**
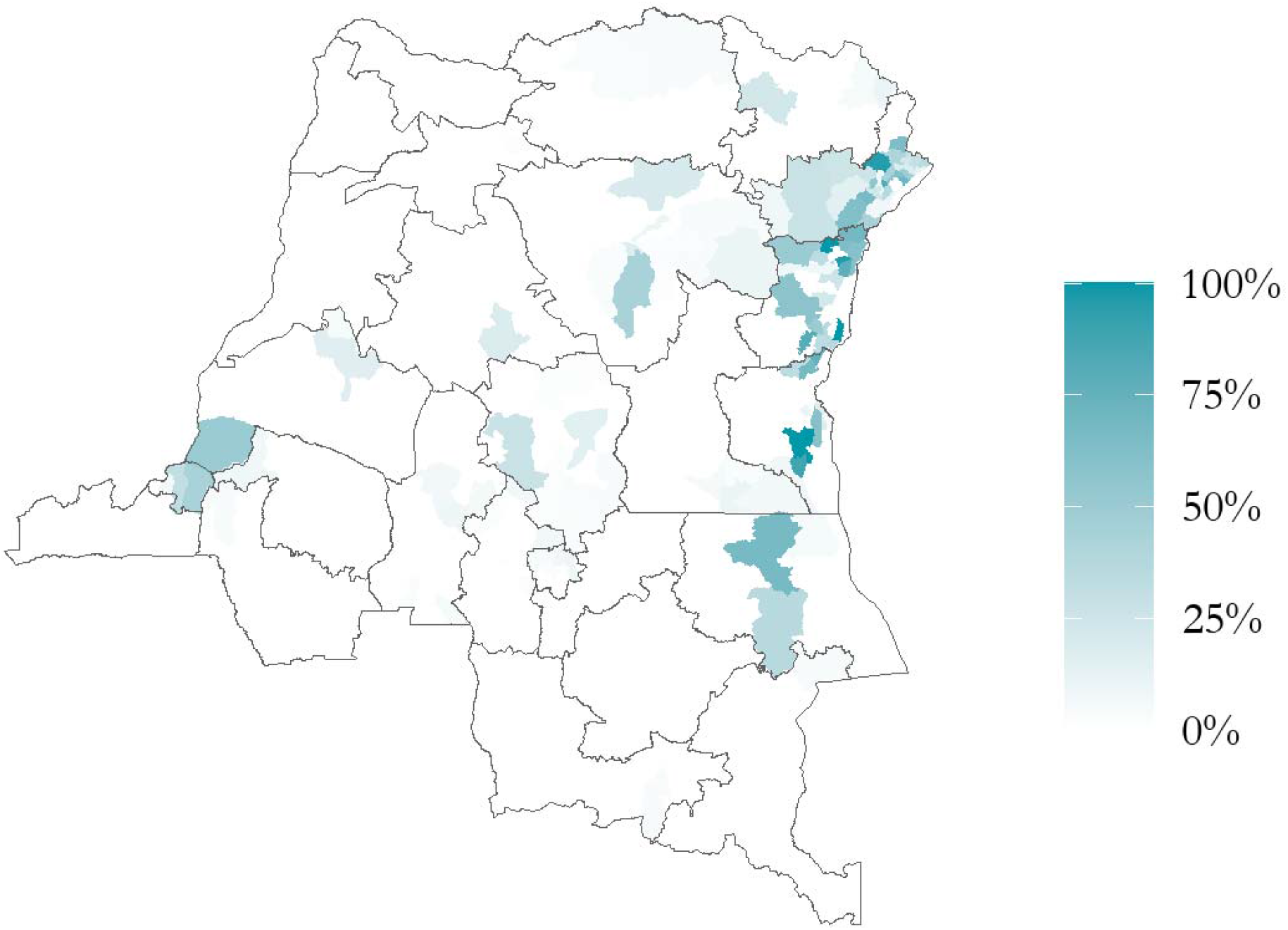
Percent of caregivers citing a response categorized as ‘War, armed conflict, ethnic conflict’ as their primary ‘other’ reason for having a zero-dose or under-immunized child.

## Discussion

Our findings align with the current scientific understanding of the capabilities and limitations of large language models (LLMs) for analyzing qualitative data. By creating a benchmark subset of the ECV dataset, we were able to rigorously test several approaches and models for categorizing free-text survey responses. The results demonstrate that NLP and LLM based approaches can achieve 61.5% to 96%, consistent with other reported performance outcomes for different workflows on similar tasks. Moreover, our study reinforces existing evidence that model performance can be further optimized when given more context or task-specific information, whether through fine-tuning or few-shot learning [27,28]. However, the amount of context and task-specific information to increase accuracy above 90% is still relatively low, enabling researchers and analysts to assign a role and provide between 20 and 100 examples for the few-shot learning approach. In our study, fine-tuning and few-shot learning can achieve similar performance, but there are differences between the computational complexity and cost including having a higher upfront cost for fine-tuning, but a potentially lower per model call cost. Overall, our study of a real-world example from the ECV surveys underscore the reliability and feasibility of LLMs in automatic large-scale qualitative analysis, particularly when enhanced by targeted, researcher-driven training strategies.

This LLM-based approach with a relatively low number of researcher-provided training examples provides the opportunity to enable new avenues of research. These include analysis of existing free-text data previously deemed too costly to work with and improving survey design flexibility and quality control. Furthermore, the ability to efficiently structure free-text data offers opportunities for researchers to rethink existing survey data paradigms.

Our study provides several novel contributions to both AI applications and global health research, particularly through substantive lessons learned from the ECV survey data. To our knowledge, this is one of the first applications of LLMs to large-scale survey data containing free-text responses in Global Health to generate novel and actionable insights into human behavior and motivations. One key insight is that our results demonstrate that AI-assisted analysis of free-text responses provides a flexible tool to track transitory or unexpected events that survey designers may not have anticipated when developing response-options in a structured questionnaire, i.e., the COVID pandemic and the health care worker strike as reasons for non vaccination identified in the survey text data. This approach also enables real-time adaptability, giving researchers the ability to identify quantitative insights from responses that fall outside pre-determined categories in the survey. Both the flexibility and quantitative components of this approach could be quite helpful for emerging public health or socio-political concerns, which might not be reflected in traditional survey designs methodology Another key insight is the realization that free-text responses, while seemingly unstructured, are still influenced by the context of the survey—especially by the interviewer. For example, responses categorized under “negligence” included examples such as ‘Laziness of the mother’ and ‘Mother was negligent’ which might reflect biases in how responses were interpreted by human interviewers. As such, while free text may represent a more raw format of data than pre-selected categories, the responses themselves may still not accurately represent the perspective of the respondents. This insight underscores the importance of complementary data collection methods, such as audio recordings, to capture a more authentic representation of respondents’ motivations. Recent advances in audio-to-text models, LLMs, and language representation offers a compelling future for survey implementation to assess key questions of behavior and motivation in global health applications.

Beyond its immediate use case, our study offers a forward-looking framework for advancing survey research. First, existing datasets containing free-text responses can be reanalyzed with LLMs to uncover previously overlooked trends. A few examples of such datasets include Demographic and Health Surveys (DHS) and Performance Monitoring for Action (PMA) surveys, which frequently contain open-ended responses. Second, future survey designs could include more open-ended questions, relying on LLMs to efficiently categorize responses during post-survey analysis. This study provides a low-cost, scalable solution for global health organizations to analyze their qualitative data with minimal researcher input while maintaining high accuracy. Additionally, the methodology outlined here offers a pathway for validating AI-generated results, making it a reliable tool for policy analysis. For example, policymakers could use these models to subset data by geography or demographic group, allowing targeted analysis of key health issues. This not only democratizes access to data but also empowers stakeholders with actionable insights for more localized decision-making in global health initiatives.

There are several key limitations and challenges to this research. One key limitation of our approach is the reliance on closed-source models like GPT-4o, where the underlying mechanisms, including the prompts and training data, are not fully transparent to the public. This opacity can make it difficult to fully assess the reasoning behind certain outputs or improve models with more transparency. However, by creating a benchmark dataset and rigorously testing LLMs against it, we can mitigate some of the risks posed by this lack of visibility. This structured validation helps ensure that the model’s categorization is reliable and interpretable, even if the underlying mechanisms remain proprietary. Another challenge is the cost associated with some of these models, which can be prohibitively expensive for organizations and institutions, especially in low-resource settings. Our study serves as a roadmap for implementers by comparing performance across different models, helping users weigh accuracy, cost, and feasibility for their specific use cases and budgets. Additionally, the computational complexity of implementing LLMs may present a barrier for some researchers or organizations. To address this, providing open-source code and resources will be crucial to democratizing access to these tools, enabling more global health stakeholders to benefit from AI-driven insights.

Another limitation in this study was the pre-definition of categories using expert knowledge and exploratory analysis. While this approach helped focus the categorization, it also constrained the ability to discover new, emergent categories that could offer novel insights. Early computational experiments revealed the sensitivity of the models to role instructions when attempting to generate new categories, suggesting the need for more refined techniques. Future work should also involve multiple human coders and consensus panels to assess both AI and human-generated categorizations, providing a more rigorous benchmark. Furthermore, continuous validation across different question domains, languages, and model versions is essential. As AI models rapidly evolve, regular testing and benchmarking will be necessary to keep the global health community informed on best practices for applying AI-driven analysis to survey data.

Despite those challenges, the integration of generative AI tools in global health data is poised to have a substantial impact. Large language models (LLMs) can categorize and summarize unstructured text data using various approaches, offering an unprecedented opportunity to scrutinize vast amounts of existing data and paving the way for innovative methods of gathering new insights. With emerging multi-modal capabilities, data collection will evolve to empower even highly capable yet under-resourced teams, enabling them to achieve meaningful results. As models improve, become more cost-effective, and open-source options come online, deploying LLMs on smaller, peripheral devices will become increasingly feasible. This will create opportunities for more accurate, timely, and localized data, which will ultimately lead to better-informed decision-making across low- and middle-income countries (LMICs). By reducing barriers to advanced data analysis, these tools will reduce barriers to producing insights from data and enhance the ability of global health stakeholders to optimize interventions, respond to emerging public health challenges, and improve outcomes for the populations they serve.

## Data Availability

Raw survey responses to the free-text question and all code for this study in both python and R are available at
https://github.com/InstituteforDiseaseModeling/AIAugmentedSurveyResponseCategorization

## Acknowledgements

This publication is based on research funded in part by the Gates Foundation, including modeling and analysis performed by the Institute for Disease Modeling at BMGF. This study uses data drawn from the DRC vaccine coverage surveys, also funded by UNICEF, USAID, and GAVI.

## Supplementary Information

**Supp. Figure 1:**
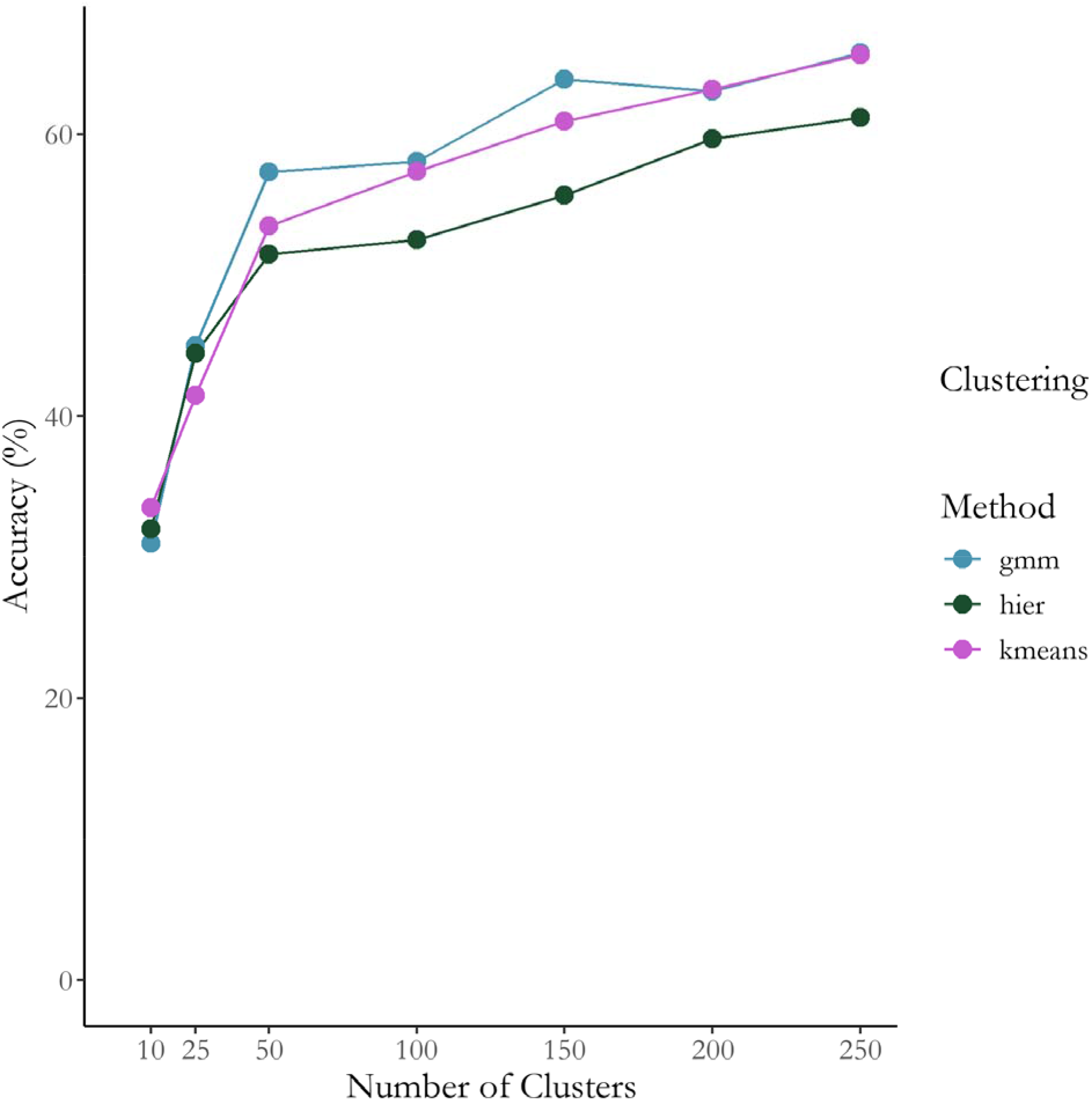
NLP-approach sensitivity to clustering choice and number of clusters.

